# Should I Stay Or Should I Go? Predicting the need for medical care after toxin exposure using SHAP-interpretable gradient boosting

**DOI:** 10.64898/2026.01.21.26344504

**Authors:** Hugo Lerogeron, Michael Chary, Laurent Gueguen, Kim An Nguyen

**Affiliations:** Hospices Civils de Lyon & Biometrics and Evolutionary Biology Laboratory, University Claude Bernard Lyon 1, Lyon, France; Department of Emergency Medicine, Weill Cornell, New York, New York, USA; Biometrics and Evolutionary Biology Laboratory, University Claude Bernard Lyon 1, Lyon, France

## Abstract

**Objective:** The most important initial step in evaluating exposure to a poisonous substance is to determine if the exposure was substantial enough to cause harm. If so, the person requires rapid evaluation by a specialist. This assessment is usually done by medical toxicologists working at poison control call centers. There is a looming knowledge gap in medical toxicology in France. Funding for poison control call centers is decreasing. No new toxicologists are being trained. Although clinical decision support algorithms exist for specific substances, there is no triage model that covers multiple substances, presenting the generalist with the problem of choosing the right tool and having no recourse for exposures where the substance is unknown. The goal of this study was to develop a generally applicable clinical decision support tool so that the general clinician can assess exposure severity in simple cases as a toxicologist would, preserving limited resources for more complex cases.

**Methods:** We evaluated the performance of decision-tree–based and gradient-boosted machine learning models to predict the triage recommendation, operationalized with a binary outcome, go to health care facility or stay at home, or a ternary outcome, urgent health care evaluation, non-urgent evaluation, and stay at home. We extracted data recorded from the Lyon Poison Control Call Center between 2000 and 2025. We included every available mono-intoxication on all patients. Cases with missing original recommendations were excluded. Missing data were left as-is. Cross-validation was used to evaluate statistical significance. We used SHAP(SHapley Additive exPlanations) to identify the most important features for predictions and assess the clinical realism of our model. Model performance was evaluated using F1-score and ROC AUC. Our benchmarks were published algorithms that focus on single-substance intoxications.

**Results:** We identified 612,569 cases and excluded 354,917, leaving 257,652 cases for model development. Recommended dispositions were: stay at home 63.3%, emergency facility 29.0%, and non-emergency facility 7.7%. The study included patients of all ages, with a median age of 9 years. The most frequent toxic exposures involved carbon monoxide inhalation and the oral ingestion of analgesics and anxiolytics. For the binary task, CatBoost achieved the best performance (F1 = 0.801; ROC AUC = 0.890). For the three-class task, LightGBM performed best (macro F1 = 0.654; multi-class ROC AUC = 0.867). The circumstance of exposure, SNOMED codes and agent were the most influential features. Our results were competitive with algorithms focusing on intoxication due to a single substance.

**Conclusion:** Gradient-boosted tree models can produce accurate, interpretable, and clinically relevant predictions of poisoning severity from routine PCC data. With external validation and prospective testing, such tools could complement expert judgment to improve triage consistency and patient outcomes.

## 1 Background and objectives

In France, when someone has been exposed to a potentially toxic substance, they or their caregiver calls a poison control center (PCC) to inquire about the level of care needed. A healthcare professional (HP), using their knowledge and experience, must then assess the severity of the intoxication. To do that, they elicit data such as their age and weight, symptoms, liver or kidney dysfunction, the type of substance and dose, and the time between exposure and the call. Despite possibly inaccurate or missing information, the HP must make the appropriate triage decision without delaying care or consuming unnecessary resources.

Making this decision is a challenging task. Any compound can be poisonous in the right amount, even water. Across developed nations, most calls to PCC involve children, 55% in France, requiring knowledge of pediatrics and toxicologists. Most cases require no intervention (63.3%), and nearly half of patients referred to the hospital for treatment of a suspected poisoning receive no treatment, even if referred by Poison Control [1], leading to the challenge of identifying cases that are rare but fatal without intervention. The volume of cases is also significant. The Lyon PCC gets 80 to 100 calls every day, representing approximately 360 to 450 human exposures per 100,000 inhabitants annually (based on the 8.1 million population of the Auvergne-Rhône-Alpes region).

To help HPs make faster, more accurate decisions, various decision-support algorithms have been proposed. Here, we lump them into the following categories: conventional approaches, rule-based reasoning, case-based reasoning, and machine learning-based reasoning. There is no standard in clinical practice for grading the severity of exposure or prognosticating the clinical course.

### 1.1 Regression Models

The Poison Severity Score (PSS) is endorsed by the International Programme on Chemical Safety, the Commission of the European Union, and the European Association of Poison Centers and Clinical Toxicologists (IPCS/EC/EAPCCT) [2, 3]. It grades the clinical severity based on observed symptoms. Although the PSS is used in research as a benchmark [3, 4, 5, 6], it contains elements that require the expertise of a toxicologist to fill out [7] and requires information that a person calling from home does not have: biological markers such as *HCO*_3_, ASAT, CPK, or the ability to grade symptoms severity (for instance, the average caller can not differenciate between mild or “standard” haemolysis).

It is technically not a prognostic score, but rather a tool to assess the severity level when clinical features are at their most critical: a higher grade will at least lead to a follow-up of the case by HPs[2].

Most models for grading the severity of a potential poisoning are geared towards determining what level of inpatient care is appropriate and assume that the patient is in a healthcare facility and has had laboratory testing.

The Tanta University Risk Model (TURM) predicts the need for critical care (also termed intensive care or level 3 care) with (AUC = 0.79, N = 293) [8]. Flack and colleagues obtained similar results (AUC = 0.87, N = 400) using the same model in an independent cohort.[9]. TURM considers the patients level of awareness via the Glasgow Coma Scale, vascular tone via the diastolic blood pressure, and acid/base status and respiratory drive via the respirator rate, oxygen saturation, and serum bicarbonte.

The ICU Requirement Score predicts whether a symptomatic poisoned patient still needs the ICU after 24 hours of inpatient care at any level [10]. It considers age, the heart rate, the systolic blood pressure, the class of ingestant, mental status, respiratory status, the presence of other life-threatening conditions, and the presence of dysrhythmias. This score was validated twice on a French dataset [11, 4]. However, this scoring system does not generalize beyond critically ill patients, and its recommendations even for critically ill patients substantially disagree with those of toxicologists [12]. Using only age, oxygen saturation, and point-of-care lactate, [13] introduced the poisoning Early Warning Score (pEWS), and obtained an area under the curve of 0.896. However, its utility is constrained in a triage context as it requires clinical data typically unavailable to a caller at home and was primarily designed for hospital-based risk stratification rather than initial pre-medical evaluation. Scores have been developed for specific intoxications, like [6] for acute poisoning with central nervous system xenobiotics, [14] for acute antipsychotic poisoning, or [15] for ethanol poisoning. The primary constraint is their high specificity: a clinician would theoretically require a validated model for every possible agent, which seems impossible considering that every substance can theoretically cause an intoxication. Moreover, the agent is unknown in some cases, highlighting the need for generalist approaches.

### 1.2 Expert Systems

Expert systems use a set of predefined rules and logical statements to make predictions. An example of such a rule could be: *If a patient’s temperature is over 39°C, then they have a high fever*. These rules are curated by human experts. The main advantage of expert models is their interpretability.

One of the first expert systems for toxicology was SETH (Système d’Expert en Toxicologie Hospitalière), developed at Rouen University Hospital [16]. Unlike simple lookup tables, SETH utilizes an inference engine to process patient-specific inputs, including the ingested drugs, doses, time since exposure, and physiological markers, against an extensive knowledge base of over 1,000 toxicological rules. A seminal demonstration of decision support, SETH was shown to significantly reduce triage errors among junior physicians by providing standardized recommendations for levels of care, ranging from home monitoring to intensive care admission [17].

The main limitations of expert models lie in the rule-making step. Experts need to make a comprehensive and internally consistent set of rules, which can be very time-consuming to create for clinically realistic scenarios and challenging to maintain. As an intoxication can be due to virtually every substance that exists, resulting in hundreds of possible clinical effects, few models have been developed.

As a result, few attempts have been made to use expert models in this context. Still, Chary et al. [18] used a probabilistic logic network (PLN) to diagnose acute poisoning. A PLN consists of pairs of probabilities and logical statements, acting as rules.

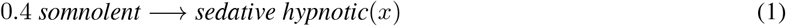

Equation 1 is an example of such pairs, showing that in 40% of cases, if a patient is somnolent, then they may have been intoxicated by an agent of the sedative hypnotic class. 34 of those pairs were used to predict the toxidrome of a generated case, showing performance similar to that of humans on easy cases but underperforming clinicians on more complex cases.

### 1.3 Machine Learning Models

The goal of machine learning (ML) methods is to learn a model that automatically maps inputs to the desired outputs. Here, it would mean feeding the model patients’ data to predict the severity of the intoxication directly. While conventional approaches and rule-based reasoning require expert knowledge for model development, ML models are complex algorithms designed without specific dependence on the actual use case. However, the model learns by trying to mimic the prediction of actual experts: their help is therefore still needed. This paradigm is called supervised learning [19]. ML approaches can leverage huge databases to learn their task, and can afterwards be used to predict for new patients. ML has been shown to outperform conventional approaches on ICU prognosis [20] and has recently been used on numerous toxin exposure predictions: [21] on acute methanol poisoning, [22] on acute organophosphate poisoning, [23] on diphenhydramine exposure, [24] on acetaminophen or [25] on acute diquat poisoning. In particular, models based on decision trees perform the best. The goal of decision trees is to learn implicit decision rules (logical rules of the form *if then*, generally comparing a variable to a threshold) from the data. By themselves, decision trees are easy to understand. In most applications, however, models are made of numerous decision trees, with each decision tree only considering a subset of the dataset, like in Random Forest. Decision trees are used as a set of weak learners, whose predictions are averaged to get a stronger, less biased prediction. When trees are trained iteratively to improve on the previous tree’s mistakes, we get gradient boosting algorithms, such as XGBoost. These advanced algorithms trade higher performance for worse interpretability similarly to deep learning models.

Deep Learning (DL)[26], a subset of machine learning, uses artificial neural networks to learn complex patterns in the data. There exist various types of standard architectures for DL models, such as convolutional neural networks, recurrent neural networks, dense neural networks, etc [27]. In the context of intoxication severity assessment, data is mostly tabular, which is still an area where deep learning models struggle to outperform algorithms based on decision trees [28]. While predicting the need for intubation for patients poisoned by methanol, [29] confirms that decision trees outperform various architectures of deep learning models.

Despite their performances, the use of ML models is limited by the difficulty of understanding their predictions. Indeed, conventional approaches are based on simple models; it is easy for an HP to understand why a prognosis prediction was made. In the same way, rule-based reasoning aims at mimicking the way HPs make a decision, and case-based reasoning justifies its prediction by the most similar known case. The explainability of those approaches is called *Ante-hoc*: HPs can directly understand the prediction [30]. On the other hand, ML algorithms are made of numerous complex mathematical operations, making their functioning akin to a ‘black box’. As a result, HPs tend to have low trust in ML-based approaches and use the system less [31]. Therefore, ML methods require *Post-hoc* explanations techniques [30]. In the context of poison prognosis, explanation is mostly based on showing the importance of features on decisions [21], [29], [25], using tools like SHAP. SHAP (SHapley Additive exPlanations) is a method that assigns each feature a numerical value representing its contribution—positive or negative—to a model’s prediction, based on principles from cooperative game theory^1^.

Approaches and the variables used are summarized in the table 1.

**Table 1.** Summary of predictive models and required clinical variables in toxicology.

| Model / Score | Category | Required Variables & Clinical Features |
| --- | --- | --- |
| PSS [2] | Clinical Grade | Observed symptoms, biological markers ($HCO_3$, ASAT, CPK). |
| TURM [8] | Regression | GCS, $O_2$ Saturation, Diastolic BP, Respiratory Rate, Serum Bicarbonate. |
| ICU Req. Score [10] | Regression | Age, Heart Rate, Systolic BP, Mental Status, Respiratory Status, Ingestant Class, Dysrhythmias. |
| pEWS [13] | Regression | Age, $O_2$ Saturation, Point-of-care Lactate. |
| SETH [16] | Expert System | Ingested substances, Dosage, Time since ingestion, Age, Weight, Symptoms. |
| PLN [18] | Probabilistic | Symptom clusters (e.g., somnolence, miosis), suspected drug class. |
| Specific ML Models [21, 25, 24] | Machine Learning | Toxin-specific lab values (e.g., Methanol levels, Creatinine, pH, WBC count, serum enzymes), GCS, and Lactate. |
| Ours | Machine Learning | **Initial phone call data:** Age, Weight, Sex, Agent, Dose, Route of exposure, and Symptoms. |

Overall, while these approaches show various ways to help HPs improve poison prognosis, they all have the same limitations, namely that they require numerous biological data and are used to predict the prognosis of a few toxins. To the best of our knowledge, no ML model currently exists that predicts poisoning severity using only information obtained during the initial phone call for every possible agent. Accordingly, this study presents an ML-based approach to predict the need for medical care, while simultaneously performing triage of minor cases in which exposed individuals can safely remain at home, using only data available during the first contact with a PCC. Our objective is to develop an algorithm capable of assessing intoxication severity across all substances represented in our dataset. We systematically evaluate multiple machine learning algorithms using comprehensive performance metrics to identify the optimal predictive approach. We highlight the most important features and the values that most influence the decision. Additionally, we benchmark our method against existing approaches that assess intoxication severity for specific subsets of toxic substances, demonstrating the broader applicability and effectiveness of our proposed solution.

## 2 Materials and methods

The goal of our model was to fulfill the following objectives :

- Accurately predict whether a patient needs medical care or not, using only the information provided during the first call to the poison center.
- Provide as much information as possible to help the practitioner understand the prediction.
- Ultimately, reduce triage error.

### 2.1 Dataset

During each call, HPs systematically recorded comprehensive patient information, including demographics, presenting symptoms, medical history, circumstances of exposure, timing and location of the incident, and, when available, the specific substance involved and its dosage. Based on this information, HPs provided recommendations to callers regarding the appropriate level of medical care required, ranging from no treatment necessary to emergency department referral. This recommendation served as our target variable for prediction. Cases lacking this critical outcome information were excluded from the analysis.

#### 2.1.1 Data preprocessing

The following section outlines the processing pipeline used to convert the dataset, extracted from Lyon’s CAP database via SQL, into a format suitable for machine learning.

First, we removed multiple entries for the same patient, keeping only the oldest record corresponding to the first call to the PCC. This approach simulated an operator receiving a call about a brand-new patient, as typically occurs in PCCs.

We then performed initial data cleaning by standardizing text fields and handling missing values. We corrected age calculations by replacing empty strings and “NaN” entries with proper null values, converting comma-separated decimals to dot notation, and removing physiologically unlikely ages (those over 125 or below 0 years). We applied similar validation weight values, removing them when outside the 0–200 kg range. We encoded the symptom information using the Systemized Nomenclature of Medicine – Clinical Terms (SNOMED[32]) terminology.

We identified the primary causative agent through a hierarchical extraction process applied to the agent detail strings. The algorithm prioritized the most specific agent classification (level 7) and moved progressively to broader categories (levels 6–1) when more detailed information was unavailable. We removed multi-intoxications (12.5% of the dataset) because this study focuses exclusively on mono-intoxications. We extracted exposure information from multi-level nested structures that contained the estimated amount, confidence in the estimation, exposure circumstances, and exposure route. We converted all temporal measurements into a standardized minute-based format. This standardization enabled consistent temporal analysis across the varied measurement units present in the original data. Finally, we standardized healthcare facility information using a two-tier or three-tier classification system. The primary categories included home/exposure site (H), emergency facilities (EF), and non-emergency healthcare facilities (NEHF). This information served as the label we aimed to predict. For binary classification tasks, we merged EF and NEHF into a single category (HF).

### 2.2 Machine learning models

Following [33] and [21], we focused our efforts on decision tree-based algorithms: Random Forest (RF), XGBoost, LightGBM, CatBoost, and HistGradientBoostingClassifier(HBC)^2^. All the models had their default hyperparameters.

### 2.3 Training and evaluation

The model development process followed a structured pipeline of data partitioning, cross-validation for optimization, and final independent testing. Initially, the dataset was split into training and test sets (with a 90/10 ratio); the latter was strictly reserved for the final performance assessment to ensure an unbiased evaluation of the model’s generalizability.

To identify the optimal model architecture and calibrate decision thresholds, we implemented 10-fold stratified cross-validation on the training data. This stratified approach was chosen to ensure that each fold maintained the same class distribution as the overall dataset, thereby preventing bias during the model selection phase. To address class imbalance, we adjusted the loss function weights during training. For each model *m* and each fold *k*, we computed the optimal decision threshold *t*_*m,k*_ maximising the macro f1-score. The final chosen threshold *T*_*m*_ was the average of the 10 aforementioned thresholds.

We used the following metrics to compare the models :

- Precision: 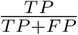
- Recall (or sensitivity) :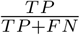
- F1 score: 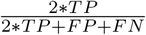
- Accuracy : 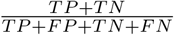
- Area Under the Receiver Operating Characteristic Curve (ROC AUC)^3^

In particular, we used the macro F1 score to account for the substantial data imbalance in our dataset, as it equally values precision and recall. This choice reflects our clinical priorities: underestimating the risk of intoxication (false negatives) could endanger patients, while recommending medical care for too many patients (false positives) could overwhelm healthcare facilities and increase waiting times, a factor shown to raise mortality [34].

Once the optimal thresholds were computed for every model via cross-validation, the models were retrained on the entire training set. The final evaluation was conducted on the previously unseen test set, using the threshold *T*_*m*_, providing a robust estimation of how the models performs on novel data and confirming that the selection process did not lead to overfitting.

### 2.4 Explanations of predictions

Being able to explain the predictions made by the algorithm will significantly enhance its usability for practitioners. Since machine learning models, and particularly gradient boosting algorithms, are complex, we used *Post-hoc* interpretation techniques. In detail, we used the importance the algorithms give to each variable to try to explain how the predictions were made. To complement this analysis, we used the SHAP library. In particular, we use the TreeSHAP algorithm^4^ and display the mean of the absolute value of SHAP. We also used SHAP values to understand which values of variables “push” towards a prediction. In particular, for a given categorical variable *V*_*c*_ and 2 classes *C*_1_ and *C*_2_, we compute the difference of the mean of SHAP values over every observation for *V*_*c*_ and each class, i.e., *D*(*V*_*c*_, *C*_1_, *C*_2_) = *mean*(*SHAP* [:, *V*_*c*_, *C*_1_]) −*mean*(*SHAP* [:, *V*_*c*_, *C*_2_]). Positive values indicate a push towards *C*_1_, and negative values towards *C*_2_. For each *V*_*c*_, we display the 3 values that push the most (i.e., with the highest |*D*| ) towards each class.

### 2.5 Comparison with other approaches

Comparing results across different models presents significant challenges. Datasets vary between studies and are often unpublished; severity definitions differ across contributions, scoring methodologies are inconsistent, and most studies focus on restricted subgroups of toxins rather than comprehensive toxin panels. Therefore, to enable meaningful comparison with other contributions, we restricted our model’s results to a specific set of agents by computing metrics exclusively on this defined subset. We also aligned the number of classification classes, comparing binary classification contributions using our binary approach and three-class contributions using our three-class approach.

## 3 Results

We conducted all experiments using Python 3.11. We kept 257,652 unique cases for our analysis. Table 2 lists every feature used by our models, along with its percentage of missing values. Because the CatBoost, RF, XGBoost, Light-GBM, and HBC classifiers can natively handle missing values, we kept those values unchanged. We also highlight the severe data imbalance: clinicians suggested NEHF for only 7.4% of patients.

**Table 2.**
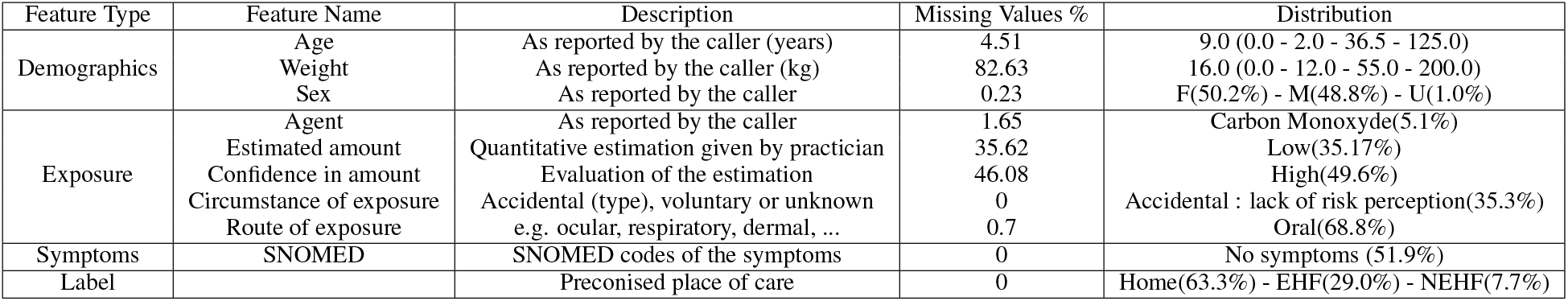
Features used by our models to predict the severity of intoxication. For numerical variables, distribution indicates the median (min - 25% - 75% - max) values. For categorical variables, we include the most frequent category. U means Unknown.

| Feature Type | Feature Name | Description | Missing Values % | Distribution |
| --- | --- | --- | --- | --- |
| Demographics | Age | As reported by the caller (years) | 4.51 | 9.0 (0.0 - 2.0 - 36.5 - 125.0) |
|  | Weight | As reported by the caller (kg) | 82.63 | 16.0 (0.0 - 12.0 - 55.0 - 200.0) |
|  | Sex | As reported by the caller | 0.23 | F(50.2%) - M(48.8%) - U(1.0%) |
| Exposure | Agent | As reported by the caller | 1.65 | Carbon Monoxide(5.1%) |
|  | Estimated amount | Quantitative estimation given by practitioner | 35.62 | Low(35.17%) |
|  | Confidence in amount | Evaluation of the estimation | 46.08 | High(49.6%) |
|  | Circumstance of exposure | Accidental (type), voluntary or unknown | 0 | Accidental : lack of risk perception(35.3%) |
|  | Route of exposure | e.g. ocular, respiratory, dermal, ... | 0.7 | Oral(68.8%) |
| Symptoms | SNOMED | SNOMED codes of the symptoms | 0 | No symptoms (51.9%) |
| Label |  | Preconised place of care | 0 | Home(63.3%) - EHF(29.0%) - NEHF(7.7%) |

### 3.1 Binary classification experiment

The results of the binary classification on the test set using balanced weights are presented in Table 3. Overall, the gradient-boosted models (XGBoost, LightGBM, HBC, CatBoost) achieved comparable performance, with CatBoost performing slightly better. A macro F1-score of 0.801 indicates a good predictive capability and suggests potential for clinical application. Furthermore, all models demonstrated near-instantaneous inference times, supporting their feasibility for real-time use. The confusion matrix is shown in Figure 1.

**Table 3.** Results of the classification distinguishing between patients needing medical care or not.

| Model | Precision | Recall | F1 | Accuracy | RocAUC |
| --- | --- | --- | --- | --- | --- |
| XGBoost | 0.791 | 0.798 | 0.794 | 0.807 | 0.879 |
| HBC | 0.779 | 0.789 | 0.783 | 0.795 | 0.874 |
| LGBM | 0.812 | 0.79 | 0.798 | 0.819 | 0.885 |
| RF | 0.761 | 0.774 | 0.765 | 0.776 | 0.855 |
| CatBoost | 0.798 | 0.805 | 0.801 | 0.814 | 0.890 |

**Figure 1.**
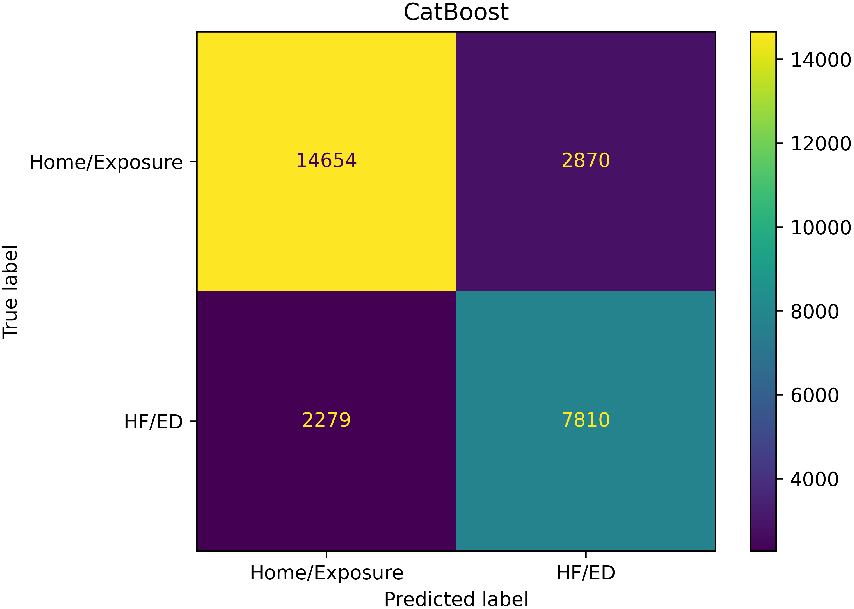
CatBoost’s confusion matrix for the binary classification task.

### 3.2 Three-class experiment

Results of the three-class experiment are given in Table 4. Once again, gradient boosting methods obtained similar performances, with CatBoost being slightly better. The confusion matrix is shown in Figure 2.

**Table 4.** Results of the classification distinguishing between patients needing emergency care, medical care, or no care at all.

| Model | Precision | Recall | F1 | Accuracy | RocAUC |
| --- | --- | --- | --- | --- | --- |
| XGBoost | 0.662 | 0.621 | 0.638 | 0.782 | 0.842 |
| HBC | 0.668 | 0.620 | 0.639 | 0.780 | 0.852 |
| LGBM | 0.667 | 0.631 | 0.646 | 0.784 | 0.856 |
| RF | 0.627 | 0.568 | 0.596 | 0.758 | 0.832 |
| CatBoost | 0.673 | 0.640 | 0.654 | 0.787 | 0.867 |

**Figure 2.**
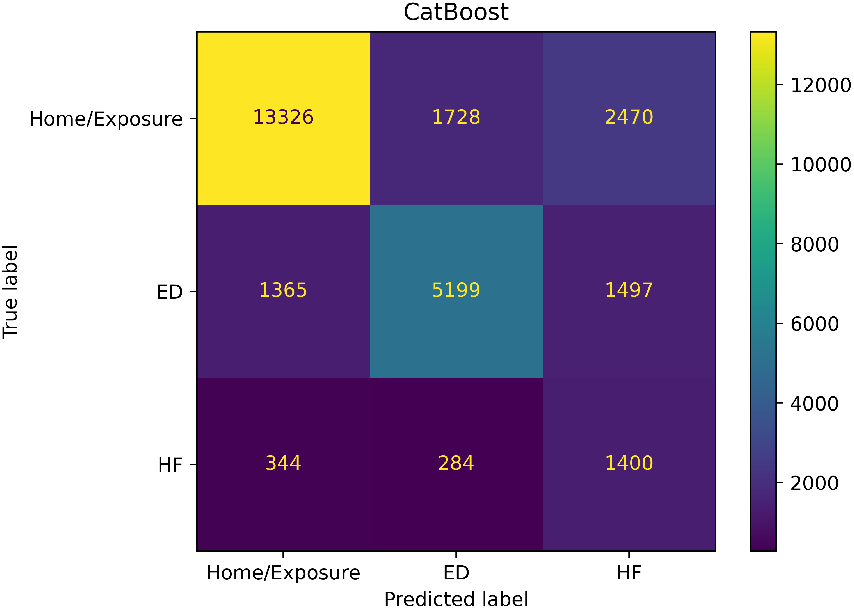
CatBoost’s confusion matrix for the 3-class task.

### 3.3 Explanations of predictions

We used the models’ built-in importance weights to highlight the most important features, as shown in Table 5 and the results obtained using SHAP, for both tasks, in Figure 3. Overall, the circumstance of exposure, SNOMED codes, and agent were found to be the three most impactful features.

**Table 5.** Importance of feature for the classification using the built-in weights.

| Feature Id | Importances |
| --- | --- |
| Circumstance of exposure | 0.2230 |
| SNOMED | 0.2093 |
| Agent | 0.1949 |
| Age | 0.1405 |
| Estimated amount | 0.1015 |
| Route of exposure | 0.0780 |
| Sex | 0.0315 |
| Weight (kg) | 0.0213 |

(b) Three-class task
| Feature Id | Importances |
| --- | --- |
| Circumstance of exposure | 0.2529 |
| SNOMED | 0.1870 |
| Agent | 0.1867 |
| Age | 0.1516 |
| Estimated amount | 0.0964 |
| Route of exposure | 0.0762 |
| Sex | 0.0322 |
| Weight (kg) | 0.0171 |

**Figure 3.**
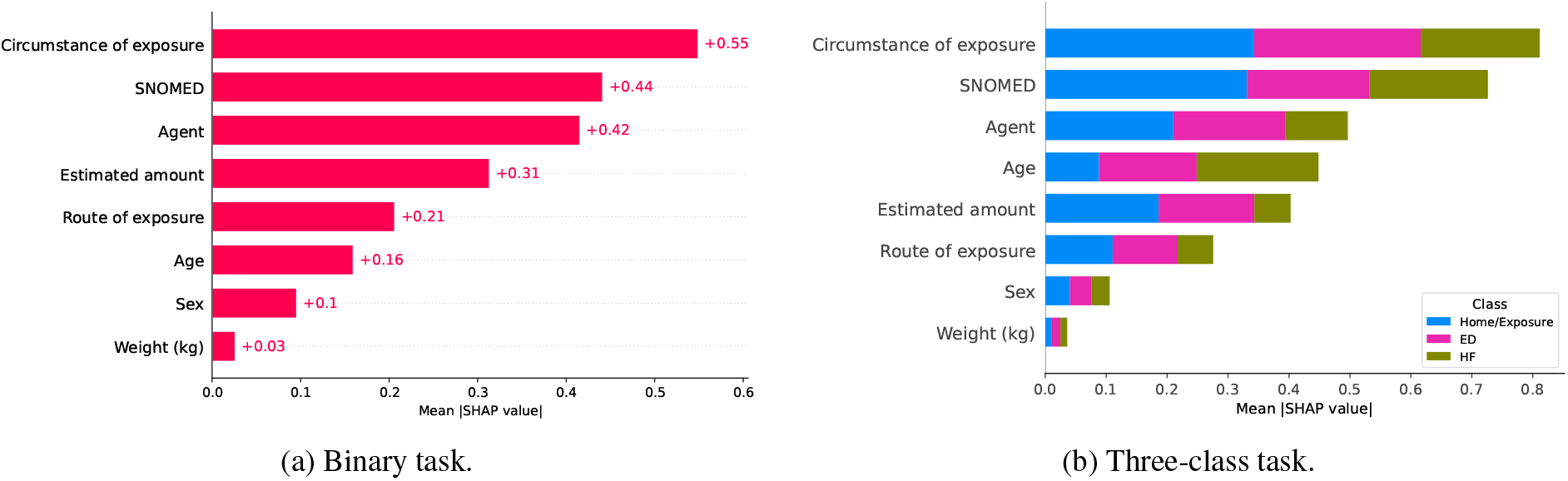
Variable importance using SHAP.

To get more insight into how the model makes its choices, we show, for each categorical variable, the 3 values that had the most impact to “push” the decision towards one class or the other. For the binary task, results are displayed in Figure 4. Among circumstances of exposure, suicide attempt and accidental undetermined were the categories most strongly pushing towards HF/ED prediction, whereas dietary, therapeutic error, and siphoning accident pushed towards Home/Exposure. Clinical features (SNOMED) showing the strongest push towards HF/ED included respiratory distress, extrapyramidal syndrome, and syncope, while ocular pruritus, nasal irritation, and distinctive taste were associated with Home/Exposure. Regarding the causative agent, Viperidae venom, terrestrial snakes, and button battery were the strongest HF/ED-pushing categories. A high or significant estimated amount pushed towards HF/ED, whereas insignificant or trace amounts and oral dissolution pushed towards Home/Exposure. Intravenous and sub-cutaneous routes of exposure were associated with HF/ED referral, while dermal, nasal, and buccal routes favored Home/Exposure prediction. Sex showed minimal discriminative contribution, with male sex exerting a marginal push towards HF/ED.

**Figure 4.**
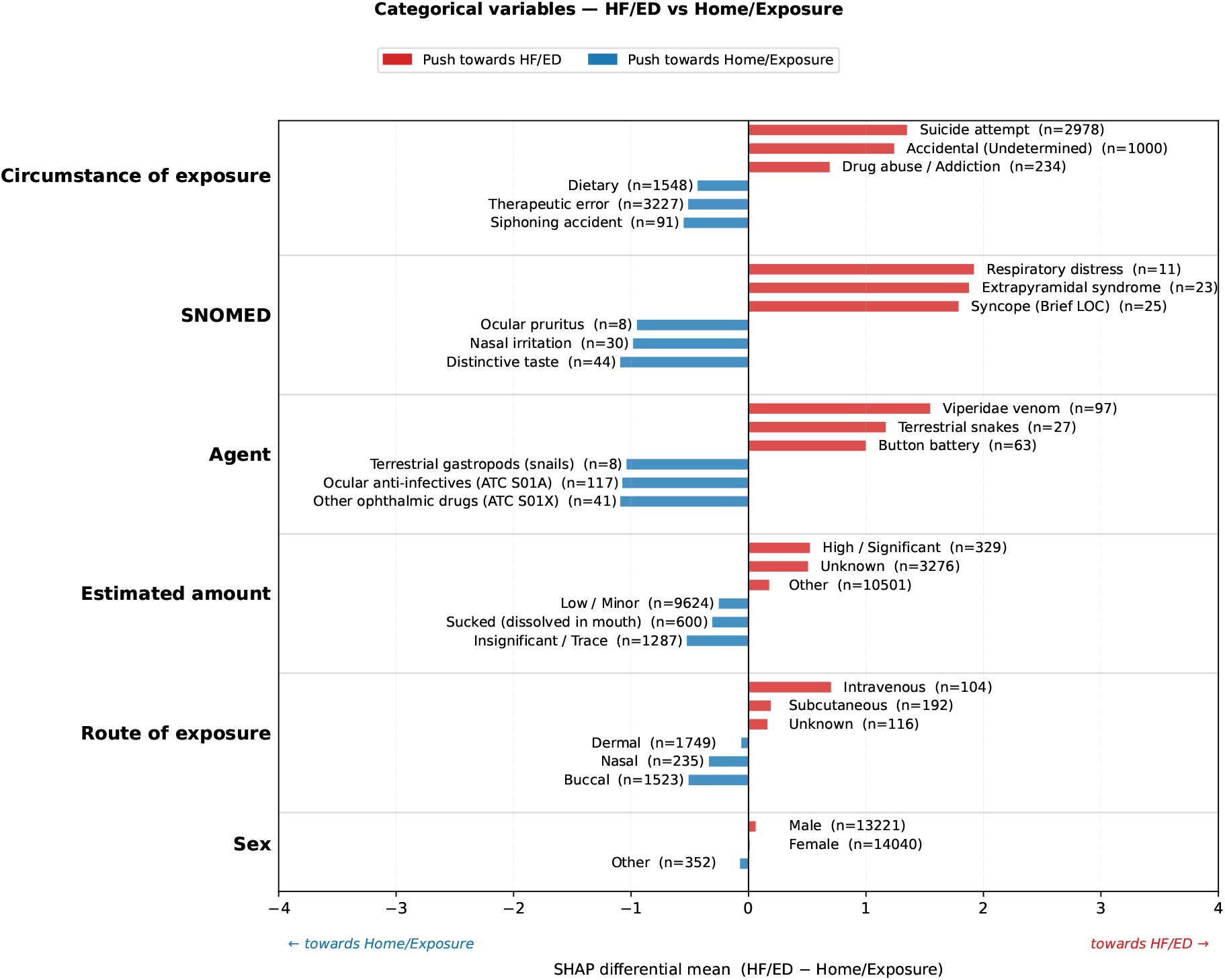
Most impactful values of each categorical variable to distinguish between home/exposure versus healthcare facilities predictions, for the binary task.

We also display the results, comparing each pair of our 3 classes in Figure 5. Suicide attempt was the circumstance most strongly and consistently pushing towards ED prediction across both ED vs. Home/Exposure and ED vs. HF comparisons, while risk perception failure and deconditioning were the circumstances most associated with Home/Exposure across both comparisons involving that class. Drug abuse pushed towards HF relative to Home/Exposure and towards ED relative to HF, suggesting these circumstances are systematically associated with higher-acuity management regardless of the specific class boundary. Among clinical features, coma at any GCS level was uniquely discriminative for ED over HF; nasal irritation and distinctive taste were recurrent Home/Exposure-pushing features across all comparisons involving that class. Regarding causative agents, Viperidae venom and terrestrial snake exposures pushed consistently towards ED across all relevant comparisons, while ocular agents and essential oils were recurrently associated with Home/Exposure. Intravenous route of exposure was the strongest and most consistent route-level predictor of ED referral across comparisons. Estimated amount and sex contributed modestly across all three classifiers, with high or significant amounts marginally favouring higher-acuity classes and sex showing negligible discriminative contribution.

**Figure 5.**
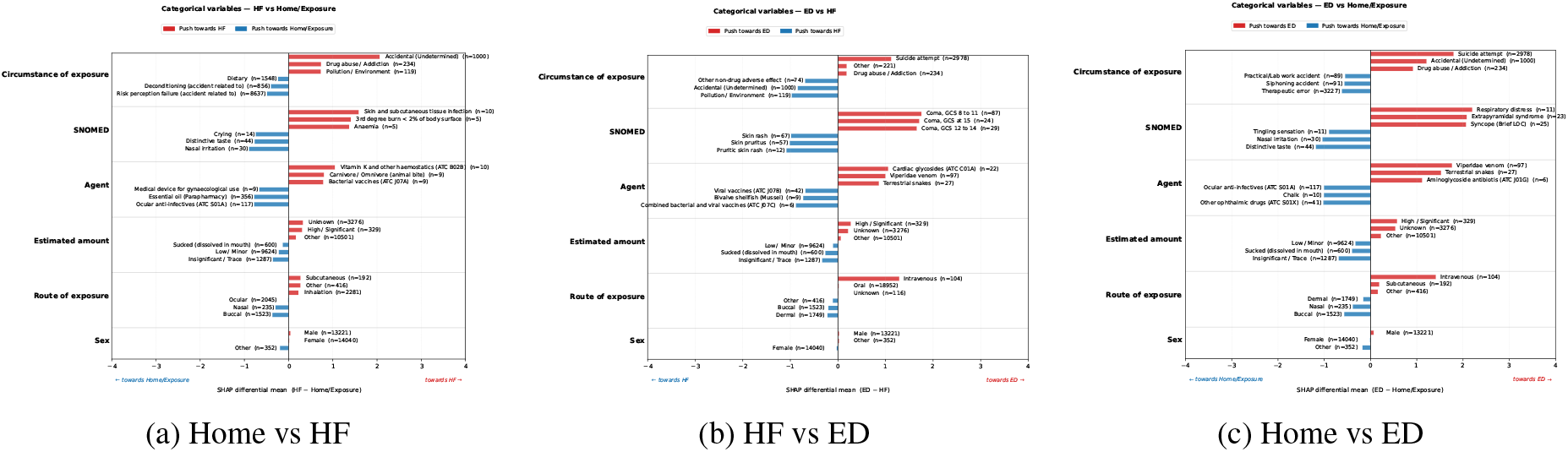
Most impactful values of each categorical variable to distinguish between classes in the ternary task.

### 3.4 Comparison with other approaches

The results are summarized in Table 6, and detailed in the following paragraphs.

**Table 6.** Comparison of toxicology prediction models with current study results.

| Study | Toxin/Target | Cases | Classes | Metric | Performance | Our Cases | Our Performance |
| --- | --- | --- | --- | --- | --- | --- | --- |
| **Two-Class Approach** |  |  |  |  |  |  |  |
| Hosseini et al. [22] | Organophosphate (mild-moderate vs severe) | 1,237 | 2 | F1-score | 0.91 | 258 (48.4% med care) | 0.77 |
| Patel et al. [35] | PICU admission | 21,646 ($\leq 6y$) | 2 | Accuracy | 0.69 | 10,223 ($\leq 6y$) | 0.84 |
|  |  | 4,085 (6-12y) |  |  | 0.53 | 1,178 (6-12y) | 0.83 |
| | | 44,683 ($\geq 12y$) | | | 0.57 | 1,748 (12-18y) | 0.85 |
| Rose et al. [36] | Carbon monoxide (mortality prediction) | 1,273 (4.5% deaths) | 2 | AUC | 0.70 | 1367 (55.3% med care) | 0.89 |
| **Three-Class Approach** |  |  |  |  |  |  |  |
| Mehrpour et al. [23] | Diphenhydramine (minor/moderate/major) | 53,671 | 3 | F1-score | 0.75 | 211 | 0.69 |
| Mehrpour et al. [24] | Paracetamol (minor/moderate/major) | 31,551 | 3 | F1-score | 0.76 | 807 | 0.62 |

#### 3.4.1 Two class approach

Hosseini et al. [22] developed a binary classification model to predict organophosphate intoxication severity, distinguishing between mild-to-moderate and severe cases in a cohort of 1,237 patients. Their optimal model employed XGBoost and achieved an F1-score of 0.91, with the most predictive features being venous blood gas pH (VBG-pH), white blood cell count, plasma cholinesterase activity, venous blood gas base excess (VBG-BE), and patient age. Given the constraints of our dataset, we could not restrict analysis to specific chemical compounds; therefore, we included all toxin categories potentially containing organophosphates: “insecticide,” “herbicide,” “vegetal pesticide,” and “animal pesticide.” Additionally, our dataset lacked access to the biological parameters that proved most valuable in their study. Our test set encompassed 258 cases, of which 48.4% required medical care, yielding an F1-score of 0.77. The most important predictive features in our model were the circumstance of exposure, the SNOMED codes, and the estimated amount of dose.

Patel et al. [35] developed a predictive model for pediatric intensive care unit (PICU) admission following acute intoxication in a cohort of 70,364 patients stratified by age. Their dataset included 21,646 patients under 6 years (14.1% requiring PICU), 4,085 patients aged 6-12 years (16% requiring PICU), and 44,683 patients over 13 years (16.7% requiring PICU). The model achieved validation accuracies of 0.69, 0.53, and 0.57 for these respective age groups. For comparison, we filtered our dataset to match these age categories and evaluated our model’s performance on predicting medical care needs. Our test set comprised 10,223 patients under 6 years (20.0% requiring medical care), 1,178 patients aged 6-12 years (24.9% requiring medical care), and 1,748 patients aged 12-18 years (67.6% requiring medical care). Our model demonstrated superior performance with accuracies of 0.84, 0.83, and 0.85 across all three age groups, respectively.

Rose et al. [36] developed a predictive model for mortality following acute carbon monoxide intoxication in a cohort of 1,273 cases, of which 57 (4.5%) resulted in death. Using a clinical scoring system based on 10 predictors (excluding biological parameters), they achieved an AUC of 0.70 on the validation set. The most important features for their model were the age of the patient, the presence of cardiac complications, and an abnormal mental status. For comparison, we restricted our dataset to carbon monoxide cases, encompassing 1367 cases with 56.1% requiring medical care. Our model achieved a superior AUC of 0.89. The five most important predictive features in our analysis were the circumstance of exposure, SNOMED codes, and the estimated amount.

#### 3.4.2 Three class approach

Mehrpour et al. [23] predicted the severity of diphenhydramine overdose using three classes: minor, moderate, and major. They constructed their dataset based on the National Poison Data System database for a total of 53,671 patients. They obtained an F1-score of 0.75, but did not report on the importance of features. For the same reasons as above, we restricted our results to “ATC R06” toxins. We had 211 cases. We obtained a macro F1-score of 0.69. The most important features were the circumstance of exposure, the SNOMED codes, and the age.

Mehrpour et al. [24] predicted the severity of acetaminophen overdose using three classes: minor, moderate, and major. They constructed their dataset based on the National Poison Data System database for a total of 31,551 patients. They obtained an F1-score of 0.76. The most important features were elevated serum levels of enzymes, other liver function test abnormalities, and anorexia. We restricted our dataset to “ATC N02” toxins and had 807 cases. We obtained a macro F1-score of 0.62. The most important features were the circumstance of exposure, SNOMED codes, and age.

## 4 Discussion

Our findings demonstrate that machine learning approaches, and particularly gradient boosted models, can accurately predict the need for emergency care after intoxications at the initial contact with the poison control center. The best performance was obtained in the binary classification experiment, which aligns with the clinical reality that distinguishing clearly benign cases from those requiring urgent evaluation is often the most critical decision. The three-class setting proved more challenging, but results still show that the model retains useful discriminative capacity.

Feature importance analysis through built-in weights and SHAP confirmed that the model’s decisions are clinically plausible. Symptoms at presentation, the circumstances of exposure, and the agent of exposure emerged as the strongest predictors. These findings are consistent with expert reasoning, providing reassurance about model validity and enhancing interpretability for healthcare professionals. In detail, the SHAP differential analysis identified feature patterns showed strong face validity with clinical expectations. The consistent association of suicide attempt with ED prediction across both ED-involving comparisons is expected, as intentional self-poisoning typically requires psychiatric evaluation and medical monitoring unavailable outside a hospital setting. The specificity of coma to the ED vs. HF discrimination is noteworthy, as it suggests that altered consciousness, while clearly driving away from home management in both cases, is a feature that further differentiates the most acute presentations requiring emergency care from those manageable in a conventional healthcare facility. Similarly, the recurrence of snake envenomation across all HF- and ED-pushing comparisons is consistent with the unpredictable systemic toxicity of viper venom, which mandates close monitoring regardless of initial severity. Conversely, the consistent association of ocular anti-infectives, essential oils, nasal irritation, and distinctive taste with Home/Exposure management reflects the typically benign and self-limiting nature of these exposures. The minimal contribution of sex to discrimination across all classifiers suggests that triage decisions in this dataset are driven primarily by clinical and circumstantial features rather than patient demographics, which is reassuring from an equity standpoint.

Compared with previous approaches relying on rule-based expert systems or conventional scoring methods such as the PSS, our model offers three main advantages: higher predictive performance across a large and heterogeneous dataset, the ability to adapt to diverse poisoning scenarios without manual rule-encoding, and transparent explanations via SHAP that may foster user trust. This balance between performance and interpretability addresses a key barrier to the adoption of machine learning in clinical toxicology.

Nevertheless, some limitations should be acknowledged. First, the dataset was restricted to single-substance exposures, whereas multi-intoxications remain common in practice and often drive the most severe cases. Second, the data come from a single centre in Lyon, which may limit generalisability to other PCCs with different populations or recording practices. Future work should therefore include multi-centre datasets and integration of multi-intoxications. Third, our algorithm obtained worse results on the non-emergency healthcare facilities class. While this could be explained by the data imbalance, it could also stem from the fact that those cases are inherently difficult to classify, even for practitioners. Biomarkers may be necessary to classify those gray-area cases better. In particular, our results show a performance gap when comparing our generalist model to specialized algorithms for the 3-class task. We define this as the ‘Generalist’s Tax’: the inherent trade-off between the breadth of coverage across thousands of substances and the high precision achievable in single-toxin models. However, the clinical utility of our tool lies in the ‘long tail’ of toxicology. While specialized models exist for the top 10 most common toxins, thousands of substances lack any validated decision support tool. Our model serves as a universal safety net, providing standardized triage logic for rare or multi-substance exposures where specialized tools are unavailable.

Regarding the choice of the target variable, while clinical outcome (final evolution) is often considered the ideal ‘ground truth’ for triage models, the Lyon PCC dataset, representative of real-world toxicology, exhibits a ‘success paradox.’ More than 99% of exposures result in full recovery, creating an extreme class imbalance that limits the discriminative utility of outcome-based labels for initial triage. Furthermore, using approaches like the PSS as a target would merely result in the model mimicking a secondary scoring system rather than the primary clinical action. Consequently, we chose HP’s recommendation as the target variable. We acknowledge that this may reproduce existing clinical biases; however, we argue that establishing a high-fidelity ‘Expert Baseline’ is a necessary first step. Before an AI can surpass human decision-making in toxicology, it must first demonstrate the ability to capture the complex, multi-factorial logic currently used by specialists to manage high-volume call centers.

From a clinical implementation perspective, the proposed model could be integrated as a real-time decision-support tool within poison control centers, assisting interns in rapidly stratifying risk and prioritizing cases requiring urgent referral. By providing an additional layer of objective, data-driven assessment, it may help standardize triage across different operators, reduce variability in advice, and ensure that high-risk patients are identified without unnecessary delay. In emergency departments, such models could also support frontline clinicians unfamiliar with toxicology by offering immediate insights into likely severity, thereby guiding monitoring intensity and resource allocation. However, successful deployment will require rigorous prospective validation, seamless integration into existing clinical workflows, and user-friendly interfaces that present model explanations clearly enough to foster clinician confidence and accountability.

## 5 Conclusion

This study shows that machine learning, and particularly gradient-boosted tree models, can provide accurate, interpretable, and clinically relevant predictions of poisoning severity from routinely collected PCC data. By highlighting the most influential variables, our approach not only supports decision-making but also reinforces clinicians’ trust in the system. With further validation and expansion, such tools could become a valuable complement to expert judgment, contributing to faster triage, more consistent advice, and ultimately improved patient outcomes in toxicology.

## Data Availability

All data are not available at the time.

## Footnotes

1 https://shap.readthedocs.io/en/latest/

2 For RF and HBC, we used their scikit-learn implementation; for others, we used their Python API

3 https://scikit-learn.org/stable/modules/generated/sklearn.metrics.roc_auc_score.html

4 https://shap.readthedocs.io/en/latest/generated/shap.TreeExplainer.html

